# ADNC-RS, a clinical-genetic risk score, predicts Alzheimer’s pathology in autopsy-confirmed Parkinson’s Disease and Dementia with Lewy Bodies

**DOI:** 10.1101/2020.04.07.20056614

**Authors:** David L Dai, Thomas F Tropea, John L Robinson, Eunran Suh, Howard Hurtig, Daniel Weintraub, Vivianna Van Deerlin, Edward B. Lee, John Q Trojanowski, Alice S Chen-Plotkin

## Abstract

Growing evidence suggests overlap between Alzheimer’s disease (AD) and Parkinson’s disease (PD) pathophysiology in a subset of patients. Indeed, 50-80% of autopsy cases with a primary clinicopathological diagnosis of Lewy body disease (LBD) – most commonly manifesting during life as PD – have concomitant amyloid-beta and tau pathology, the defining pathologies of AD. Here we evaluated common genetic variants in genome-wide association with AD as predictors of concomitant AD pathology in the brains of people with a primary clinicopathological diagnosis of PD or Dementia with Lewy Bodies (DLB), diseases both characterized by neuronal Lewy bodies. 208 consecutive autopsy-confirmed cases of PD or DLB were assessed for AD neuropathological change (ADNC), and these same cases were genotyped at 20 single nucleotide polymorphisms (SNPs) found by genome-wide association study to associate with risk for AD. In a training set of the first 127 individuals, we developed a logistic regression model predicting the presence of ADNC, using backward stepwise regression for model selection and 10-fold cross-validation to estimate performance. We then assessed model performance in a separate test set of the next 81 individuals. The best-fit model generated a risk score for ADNC (ADNC-RS) based on age at disease onset and genotype at three SNPs (*APOE, BIN1*, and *SORL1* loci). In the training set, the area under the receiver operating curve (AUC) for this model was 0.751. In the test set, the AUC was 0.781. Individuals with ADNC-RS in the top quintile had four-fold likelihood of having AD pathology at autopsy compared with those in each of the lowest two quintiles. In patients with autopsy-confirmed PD or DLB a simple model incorporating three AD-risk SNPs and age at disease onset substantially enriches for concomitant AD pathology at autopsy, with implications for identifying LBD patients in which targeting amyloid-beta or tau is a therapeutic strategy.

## INTRODUCTION

Alzheimer’s (AD) and Parkinson’s diseases (PD) are the two most common neurodegenerative disorders, together affecting >6 million individuals worldwide [9, 21]. AD is defined neuropathologically by the presence of amyloid-beta (Aβ) plaques and tau neurofibrillary tangles (NFT), while PD is defined by the presence of Lewy bodies composed of alpha-synuclein (aSyn). The average age of a patient receiving an AD clinical diagnosis is ~80 years old [2], while the average age of a patient receiving a PD clinical diagnosis is ~60 years old [27]. PD is not the only disease defined by aSyn Lewy bodies. Rather, PD belongs to a group of “synucleinopathies” collectively called the Lewy body diseases (LBD). The LBD comprise PD, with or without dementia, dementia with Lewy bodies (DLB), and multiple system atrophy (MSA) [16], with the first two entities (PD and DLB) demonstrating neuronal aSyn Lewy bodies, while MSA shows aSyn inclusions in glia. Importantly, the distinction between DLB and PD with dementia (PDD) is clinical, based on the timing of development of dementia [24]. On neuropathological examination, DLB and PDD patients are nearly indistinguishable. Furthermore, DLB and PDD share preclinical features, and shared genetic variants confer an increased risk in both disorders [3, 13, 22].

Despite traditional separation between AD and the LBD, growing evidence suggests a dynamic interaction between their pathophysiologies. Fifty to 80% of patients with a primary clinicopathological diagnosis of LBD have concomitant Aβ and tau pathology [29]. At autopsy, up to 40% of PD patients exhibit enough Aβ and NFT to qualify for a secondary diagnosis of AD [13]. Mechanistically, *in vitro* and *in vivo* studies suggest that alpha-synuclein, tau, and amyloid-beta may interact synergistically in events leading to disease development [4, 35]. From a practical viewpoint, these findings suggest that LBD patients may be at-risk for developing AD.

Genetic risk factors for developing AD have been identified through family studies and genome-wide association studies (GWAS). In a recent AD GWAS comparing >50,000 cases with >100,000 controls, 25 distinct loci were associated with risk for AD [18]. However, the genetic heritability (h^2^) reported for this study was only 0.071, and various genetic risk scores composed of AD GWAS-nominated variants have poor predictive value for AD in the general population [10].

We reasoned that the high prevalence of AD within the LBD population might enhance the ability of AD genetic risk variants to predict the development of AD pathology. Accordingly, we genotyped all common genetic variants reported in two or more AD GWAS studies to associate with AD risk in a cohort of 208 consecutive cases with a primary clinicopathological diagnosis of either PD or DLB. We then tested these AD risk variants for their ability to predict concomitant AD pathology in these cases.

## MATERIALS AND METHODS

### Participants

Clinical and neuropathological data from all autopsy cases enrolled between February 1985 and July 2019 at the University of Pennsylvania Center for Neurodegenerative Disease Research (CNDR) brain bank were assessed [36]. Those with (1) a primary clinicopathological diagnosis of PD or DLB and (2) DNA available for genetic studies were included in the analysis. All cases included in this study had a clinicopathological diagnosis of DLB or PD with or without dementia; we excluded MSA to focus on primary neuronal synucleinopathies [15]. Of 1922 accessioned cases, 208 cases met the above criteria.

Prior to conducting these studies approval was obtained from the Institutional Review Board of University of Pennsylvania and informed consent was obtained from all participants prior to death. All procedures in these studies adhere to the tenets of the Declaration of Helsinki.

### Immunohistochemistry and Neuropathological Staging

Neuropathological characterization of sixteen brain regions was conducted on all cases as previously described [1, 36]. Briefly, each region was assigned a semi-quantitative score (none, rare, mild, moderate, or severe) for tau, Aβ, and aSyn pathologies. An AD Neuropathologic Change (ADNC) score was also assigned in accordance with the National Institute on Aging’s guidelines for the neuropathologic assessment of AD. Absence of AD co-pathology was defined by an ADNC of None or Low, while presence of AD co-pathology was defined by an ADNC score of Intermediate or High [25].

### Genotyping of AD Risk Variants

Single nucleotide polymorphisms (SNPs) in genome-wide association with AD risk were nominated from the literature. Three AD genome-wide association studies (GWAS) together examining >70,000 AD subjects and >380,000 controls were used to identify candidate SNPs [14, 18, 19]. SNPs reaching genome-wide significance (p < 5 × 10^−8^) in at least two of these three major GWAS were included in our study. Twenty independent loci reached criteria for inclusion.

SNP genotype was determined by Illumina Global Screening Arrays (GSA), or TaqMan SNP Genotyping Assays, as previously described [6]. In some cases, proxy SNPs (D’>0.8 in the EUR reference population from 1000 Genomes Project Phase 3 [32]) were substituted, as indicated in the text.

### Association of Individuals Risk Variants with ADNC

Logistic regression models were used to test for association between genotype at each SNP and the presence or absence of AD co-pathology in the full analysis cohort (N=208). Because the *APOE* locus has three alleles reported to have differential effects in AD [12, 30], we considered the number of *APOE* E2 and *APOE* E4 alleles separately. Additional analyses were performed with sex and age at disease onset as covariates in the logistic regression.

### Logistic Regression Model Predicting ADNC

Autopsy cases were split into training (N=127, 61%) and test (N=81, 39%) sets, which is within the range of optimal allocation proportions for large data sets with high data accuracy [7]. The training set comprised the first batch of 127 cases genotyped, while the test set comprised the next 81 cases genotyped, with no overlap between training and test sets. Backwards step-wise regression was used to develop a binary classifier to predict the presence or absence of AD co-pathology in the training set, with age at disease onset and sex as covariates in the model. Comparison of Akaike information criterion (AIC) at each step was used to determine model fitness, and we estimated predictive performance by ten-fold cross-validation (100 iterations). The Hosmer and Lemeshow goodness-of-fit test [11] was used to evaluate the final logistic regression model developed in the training set.

Model performance at prediction of AD co-pathology was assessed in both the training and test sets using receiver operating characteristic (ROC) curves, generating an area under the curve (AUC) for both the training and test sets.

### Development of the ADNC-RS

An ADNC Risk Score (ADNC-RS) was calculated for each case based on the best model developed in the training set by multiplying the age at disease onset or the risk allele dose by the respective regression coefficient. The risk score can be used to calculate the probability of AD copathology using the formula: p = *e*^*(Risk Score) / (1 - e^(Risk Score))*. This score was generated using the “predict” function in the “caret” package in R [17] from the logistic regression model.

### Additional Details Regarding Statistical Analysis

Analyses were conducted in R (http://www.r-project.org) and Prism 8 (http://www.graphpad.com/scientific-software/prism); R-scripts are available in the Supplementary Methods. The “caret” package was used for cross-validation and model generation [23]. The “ROCR” and “pROC” packages were used for creating and analyzing receiver operating characteristic (ROC) curves [28, 31]. T-test, Wilcoxon rank-sum, or Fisher’s exact tests were used to assess differences between clinical variables, as indicated by the distributions of data. For all statistical tests, power was set at 0.8, alpha was set to 0.05, and all tests were two-sided.

## RESULTS

### LBD Cohort Characteristics

Two hundred and eight participants with a primary clinicopathological diagnosis of PD or DLB were included in this analysis. The mean age at clinical disease onset was 64.49 years (SEM 0.70) and at death was 77.67 years (SEM 0.55). The majority of these subjects (n=163/208 (78.4%)) were clinically diagnosed with PD; 108 of these PD individuals had dementia at the time of death, and 55 did not. Additional diagnoses for this cohort, as well as clinical and demographic details, are shown in **Tables 1 and 2**.

**Table 1.**
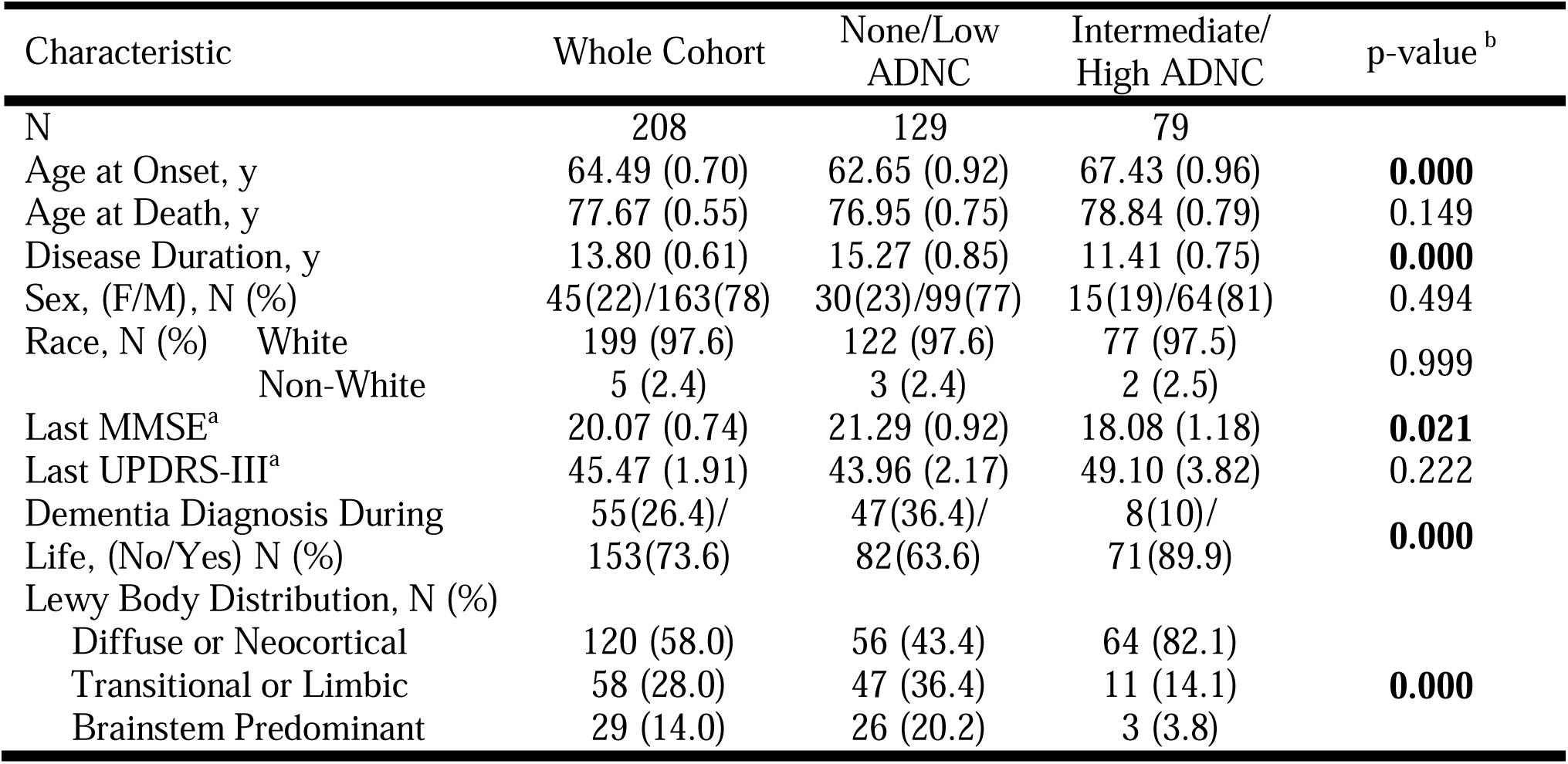
Demographics and Clinical Characteristics of Cohort. Data represent mean (SD) unless otherwise noted. ^a^Available Data (N=100 MMSE, N=68 UPDRS-III). ^b^Comparison between None/Low ADNC and Intermediate/High ADNC groups. *p<0.05.

| Characteristic | Whole Cohort | None/Low ADNC | Intermediate/High ADNC | p-value <sup>b</sup> |
| --- | --- | --- | --- | --- |
| N | 208 | 129 | 79 |  |
| Age at Onset, y | 64.49 (0.70) | 62.65 (0.92) | 67.43 (0.96) | **0.000** |
| Age at Death, y | 77.67 (0.55) | 76.95 (0.75) | 78.84 (0.79) | 0.149 |
| Disease Duration, y | 13.80 (0.61) | 15.27 (0.85) | 11.41 (0.75) | **0.000** |
| Sex, (F/M), N (%) | 45(22)/163(78) | 30(23)/99(77) | 15(19)/64(81) | 0.494 |
| Race, N (%) |  |  |  |  |
| White | 199 (97.6) | 122 (97.6) | 77 (97.5) |  |
| Non-White | 5 (2.4) | 3 (2.4) | 2 (2.5) | 0.999 |
| Last MMSE<sup>a</sup> | 20.07 (0.74) | 21.29 (0.92) | 18.08 (1.18) | **0.021** |
| Last UPDRS-III<sup>a</sup> | 45.47 (1.91) | 43.96 (2.17) | 49.10 (3.82) | 0.222 |
| Dementia Diagnosis During Life, (No/Yes) N (%) | 55(26.4)/153(73.6) | 47(36.4)/82(63.6) | 8(10)/71(89.9) | **0.000** |
| Lewy Body Distribution, N (%) |  |  |  |  |
| Diffuse or Neocortical | 120 (58.0) | 56 (43.4) | 64 (82.1) |  |
| Transitional or Limbic | 58 (28.0) | 47 (36.4) | 11 (14.1) | **0.000** |
| Brainstem Predominant | 29 (14.0) | 26 (20.2) | 3 (3.8) |  |

**Table 2.** Clinicopathological Diagnosis of Cohort. ^a^Data represent N, (%) of whole cohort. ^b^Data represent N, (%) of ADNC category.

| Clinicopathological Diagnosis | Whole Cohort<sup>a</sup> | None/Low ADNC<sup>b</sup> | Intermediate/High ADNC<sup>b</sup> |
| --- | --- | --- | --- |
| Parkinson's disease | 55 (26.44) | 47 (85.45) | 8 (14.55) |
| Parkinson's disease dementia | 108 (51.92) | 62 (57.41) | 46 (42.59) |
| Dementia with Lewy bodies | 45 (21.63) | 20 (44.44) | 25 (55.55) |

Only 43/208 (20.67%) of this LBD cohort had no ADNC at autopsy, while more than one-third had intermediate or high levels of ADNC (**Figure 1A**). Among the group with no ADNC, 16.8% are identified as Primary Age Related Tauopathy (PART). Representative immunohistochemical sections of anterior cingulate and middle frontal cortex demonstrating Lewy body pathology with co-occurring Aβ and tau NFT are shown in **Figure 1B**.

**Figure 1.**
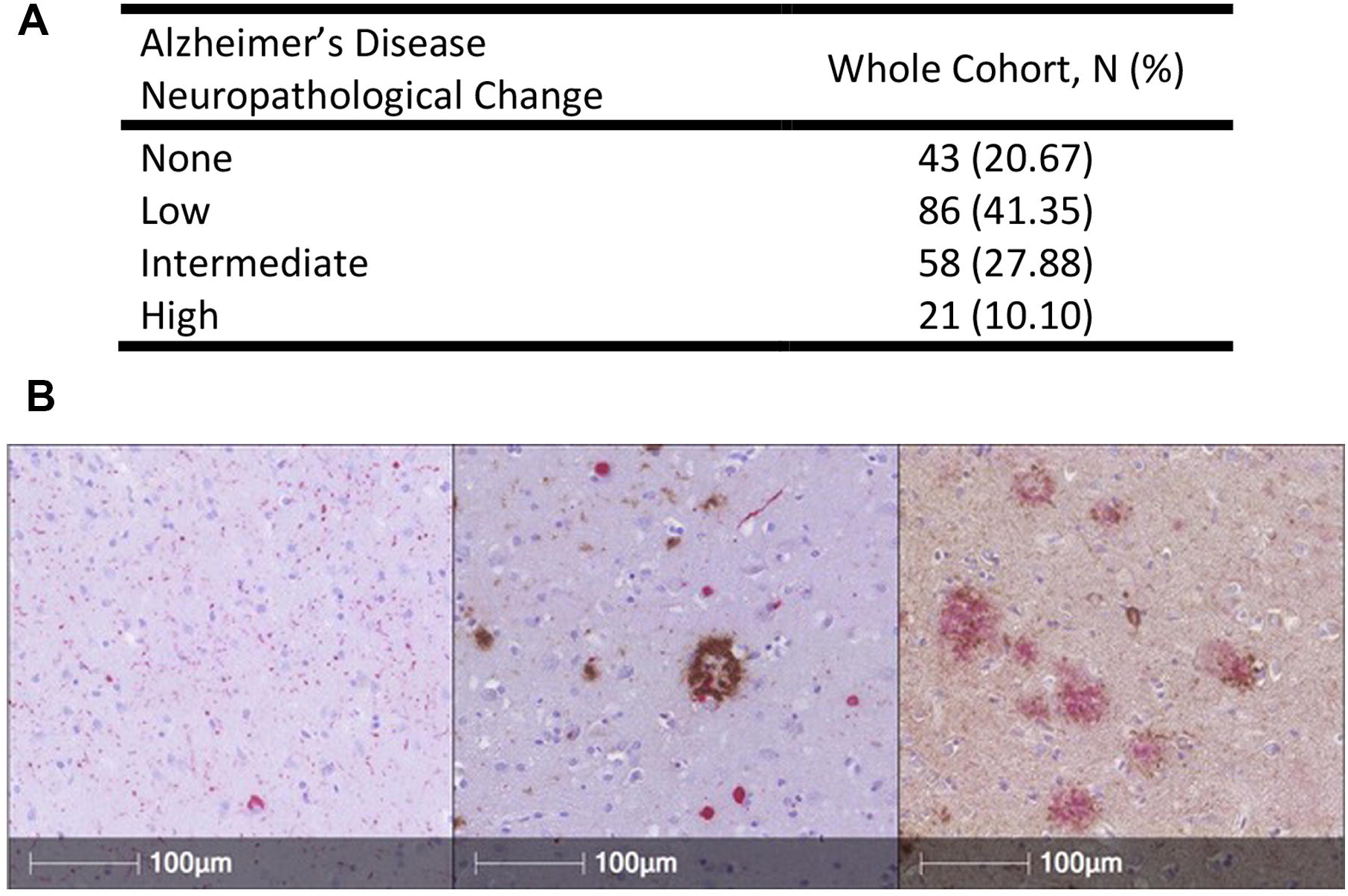
Alzheimer’s Disease Neuropathological Change (ADNC) scores in N = 208 cases with a primary clinicopathological diagnosis of Lewy body disease. A. Number of subjects and % of whole cohort at each level of ADNC. B. Representative immunohistochemical sections (160X) demonstrating Lewy body aSyn pathology alone in anterior cingulate (left panel, in red), concomitant Aβ (brown) and aSyn (red) pathology in anterior cingulate (middle panel), and Aβ (red) and tau NFT (brown) pathology in middle frontal cortex (right panel). aSyn pathology was detected with the MJFR13 antibody against phosphorylated aSyn. Aβ pathology was detected with the NAB228 antibody, and tau NFT’s were detected with the 17028 anti-tau antibody.

### Clinical Differences in PD/DLB patients with vs. without ADNC

Compared to PD/DLB subjects with absent or low levels of ADNC, subjects with intermediate-to-high levels of ADNC were older at disease onset (67.43 vs. 62.65 years, p<0.001) and had shorter disease duration (11.41 vs. 15.27 years, p<0.001). They were also more cognitively impaired, with lower MMSE scores (18.08 vs. 21.29, p=0.02), and greater rates of clinical dementia (89.9% vs. 63.6%, p<0.001, **Table 1**), prior to death. The mean time between the last MMSE and death was 2.66 years (SD 3.36).

### Association of Individual AD Risk SNPs with ADNC in PD/DLB patients

Twenty genetic loci have been robustly associated with risk for developing AD by multiple GWAS [14, 18, 19] (**Table 3**). As shown in **Table 4**, the number of *APOE* E4 alleles associated with increased risk for ADNC in PD/DLB (nominal p<0.001). One other locus near *SORL1*, represented by rs11218343, approached but did not meet the significance threshold for association with ADNC (nominal p=0.06). Adjusting for age at onset and sex minimally affected these results (**Supplementary Table 1**).

**Table 3.**
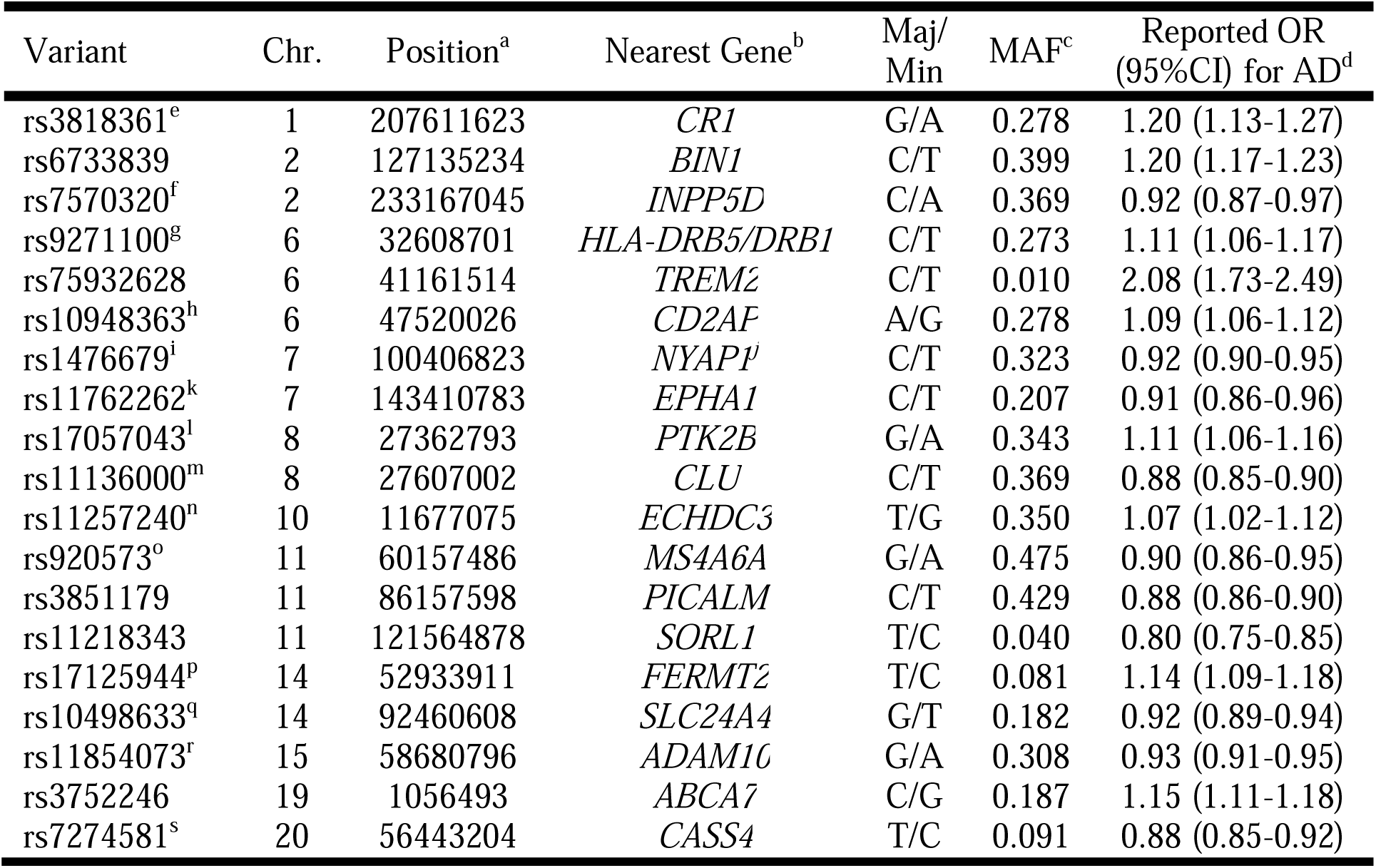
Genetic loci nominated from Alzheimer’s disease GWAS literature. ^a^GRCh38. ^b^Based on position of top SNP in reference to RefSeq assembly. ^c^1000 Genomes, CEU population. ^d^Reported by Kunkle et al (2019). ^e^proxy rs4844610 (D’=1.0), ^f^proxy rs10933431 (D’=0.84), ^g^proxy rs9271058 (D’=1.0), ^h^proxy rs9473117 (D’=1.0), ^i^proxy rs12539172 (D’=0.98), ^j^Previously the *ZCWPW1* locus, ^k^proxy rs10808026 (D’=1.0), ^l^proxy for rs73223431 (D’=1.0), ^m^proxy rs9331896 (D’=1.0), ^n^proxy rs7920721 (D’=0.95), °proxy rs7933202 (D’=0.94), ^p^proxy rs17125924, ^q^proxy rs12881735 (D’=1.0), ^r^proxy rs593742 (D’=0.88), ^s^proxy rs6024870 (D’=0.94).

**Table 4.**
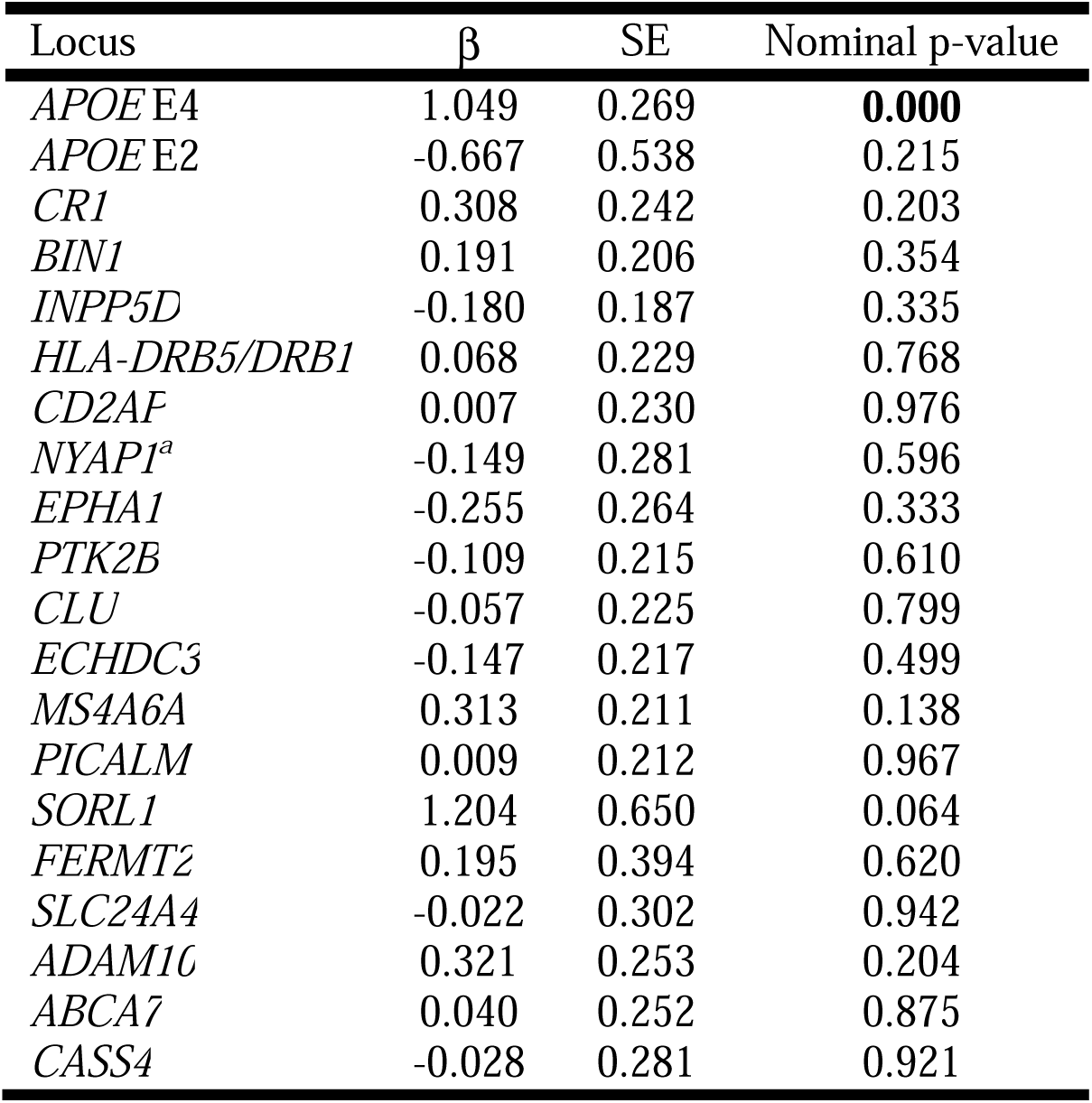
Associations between individual genetic loci and presence of concomitant AD pathology. Logistic regression coefficients (β), standard error (SE), and nominal p-values are shown. No covariates are included. *TREM2* is omitted from analysis due to lack of genetic variation at this SNP in the training set. ^a^Previously the *ZCWPW1* locus. *p<0.05.

### Development of a Model Predicting Concomitant AD Pathology in PD/DLB Individuals

In the training set consisting of the first 127 PD/DLB individuals genotyped, we developed a logistic regression model to predict concomitant AD pathology (defined as intermediate-to-high levels of ADNC). We began by including genotype at all 20 AD risk SNPs [14, 18, 19] (**Table 3**), age at disease onset, and sex in the model. We then used backward stepwise regression, with model selection based on the Akaike Information Criterion (AIC). For each model, we also estimated predictive performance by ten-fold cross-validation (100 iterations) within the training set (**Figure 2A**).

**Figure 2.**
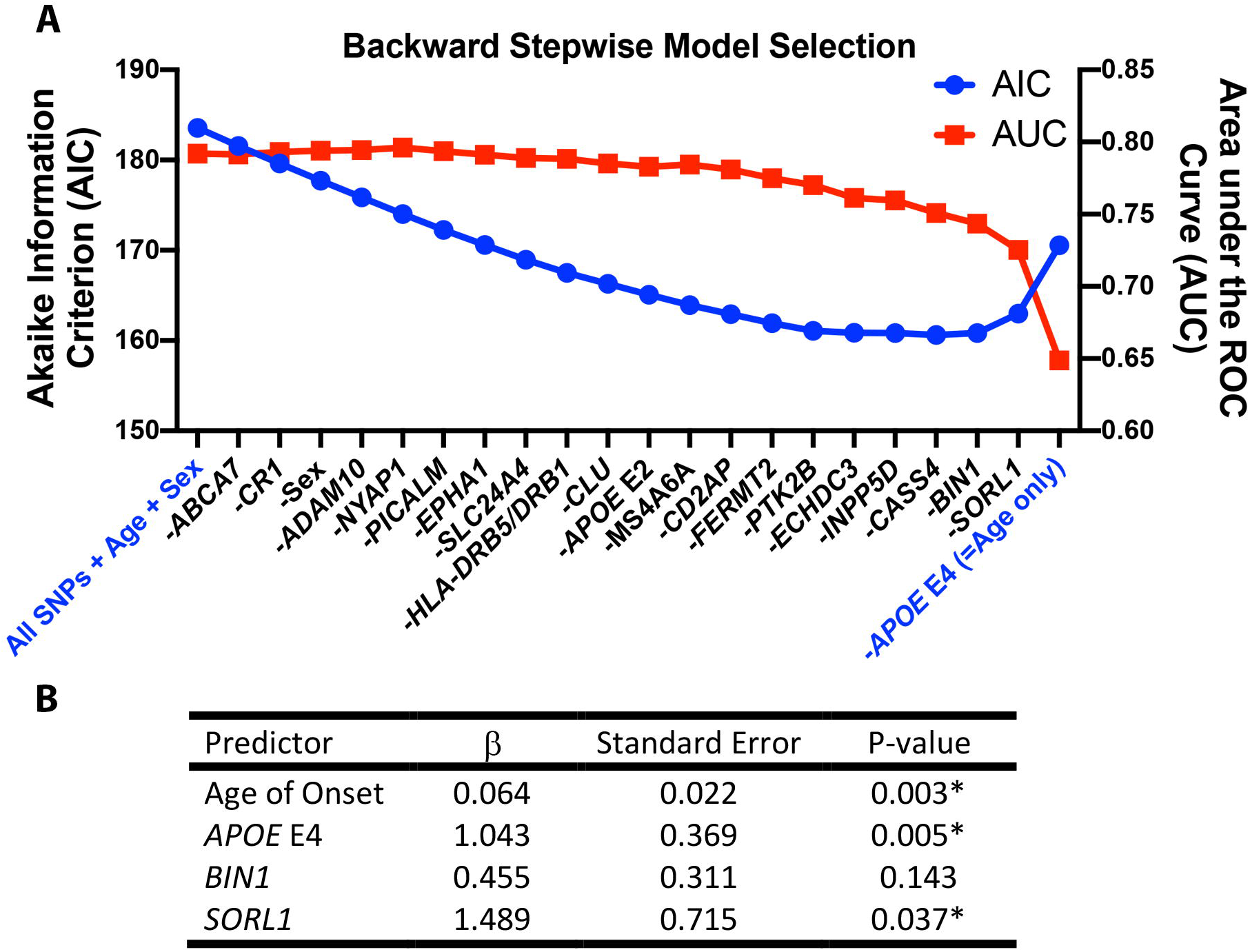
Backward stepwise logistic regression model selection for predicting concomitant Alzheimer’s Disease (AD) pathology in N = 127 cases (Training Set) with a clinicopathologic diagnosis of Lewy body disease. Concomitant AD pathology is defined as an AD Neuropathological Change (ADNC) score of Intermediate or High. A. Akaike information criterion (AIC, left axis) at each step during model selection and the corresponding area under the receiver operating characteristics curve (AUC, right axis), estimated by ten-fold cross-validation, within the Training Set are shown. Initial model included all AD risk SNPs, sex, and age as predictors; sequential elimination of predictors and effect on AIC and AUC are shown from left to right. As the Training Set cases showed no genetic variability at the *TREM2* locus, this locus was not included in the model. B. Coefficients (β), standard error (SE), and p-values for the four predictors included in the best model (lowest AIC) for predicting concomitant AD in LBD cases.

Our best model (by AIC) incorporated only four predictors: age at disease onset, number of *APOE* E4 alleles, and genotype at the *BIN1* and *SORL1* loci (**Figure 2B**). The Hosmer-Lemeshow goodness-of-fit test for this model produced a χ^2^(8, N = 127) = 7.578, p = 0.4758, indicating fit. The area under the receiver operator curve (AUC) for this model in our training set data (ten-fold cross-validation) was 0.751 (**Figure 3A**).

**Figure 3.**
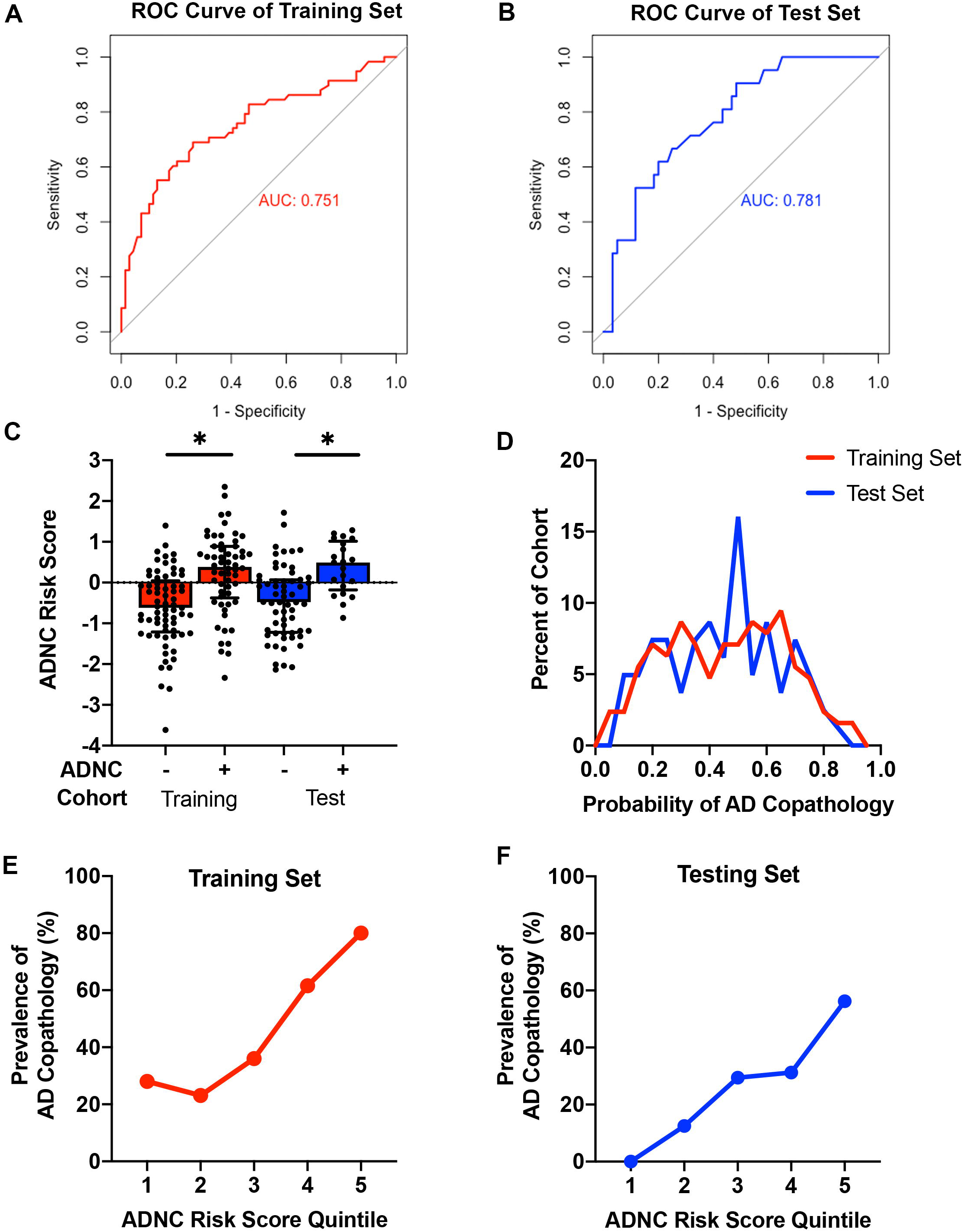
Performance characteristics of the best model for predicting concomitant Alzheimer’s disease (AD) pathology among cases with a clinicopathological diagnosis of Lewy body disease. Receiver operating characteristics (ROC) curves and areas under the curve (AUC) of the final model (with age of onset, number of *APOE4* alleles, *BIN1* genotype, and *SORL1* genotype as predictors) in the Training (A) and Testing (B) cohorts are shown. C. The Alzheimer’s Disease Neuropathological Change Risk Score (ADNC-RS) calculated from the best logistic regression model is shown in both the Training Set and Test Set cohorts. Individuals positive for ADNC showed higher average ADNC-RS. D. The probability of concomitant AD pathology was calculated from the ADNC Risk Score for each case. Values above 0.5 have a high probability of concomitant AD pathology, while values below 0.5 have a low probability of concomitant AD pathology. The prevalence of concomitant AD pathology at each quintile of ADNC Risk Score in the Training (E) and Testing (F) cohorts demonstrates fourfold enrichment for the presence of ADNC for individuals in the top quintile vs. individuals in the first two quintiles of risk. *p<0.05.

### Model Performance in Test Set

We applied the best model developed in our training set to a test set of 81 PD/DLB individuals whose data were never used to develop the predictor. Despite differences in the proportion of cases with concomitant AD pathology in the training set (46%) vs. the test set (26%), our model performed equally well in the test set, with an AUC of 0.781 (**Figure 3B**).

We additionally performed a subgroup analysis, applying our predictor only in cases with a clinical diagnosis of PD or PDD (N=163), which minimally affected the results (AUC = 0.728, **Supplemental Figure 2**).

### Development of an ADNC Risk Score

In order to develop a clinically-useful tool, we used our logistic regression model to generate a risk score for concomitant AD pathology (ADNC risk score, or ADNC-RS). An ADNC-RS was calculated for each case using the following formula:

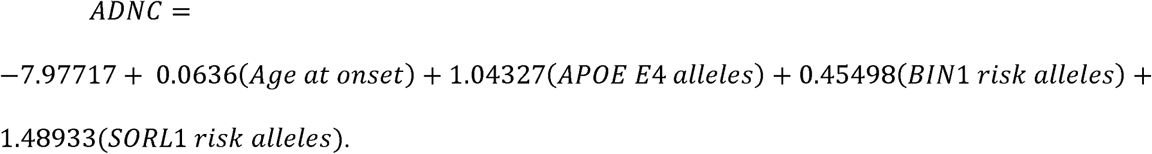

The distribution of ADNC-RS across both the training and test sets is shown in **Figure 3C**. The ADNC-RS was significantly higher for PD/DLB individuals with concomitant AD pathology in both the training (M 0.241 (SD 0.129) vs. M −0.622 (SD 0.111), t(125)=-5.120, p<0.001) and test sets (M 0.378 (SD 0.139) vs. M −0.470 (SD 0.113), t(79)=-4.061, p<0.001). For each case the ADNC-RS was used to determine the probability AD co-pathology. The distribution of ADNC-RS predicted probability of AD co-pathology is shown in **Figure 3D**. Importantly, individuals with ADNC-RS in the highest quintile were four times more likely to have AD pathology than individuals with ADNC-RS in the lowest two quintiles (**Figure 3E-F)**.

## DISCUSSION

In this study, we investigated 208 PD/DLB cases in order to determine whether common genetic variants associated with risk for AD by GWAS might predict which individuals will develop concomitant AD pathology. We first demonstrated that concomitant AD pathology is highly prevalent in PD/DLB patients, with over one-third of this group exhibiting intermediate-to-high levels of ADNC. We next evaluated a set of 20 common genetic variants found by multiple AD GWAS to associate with risk for AD, examining their association with ADNC in PD/DLB and developing a best-fit logistic regression model predicting the presence of intermediate-to-high ADNC in these primary neuronal synucleinopathies. A predictor incorporating only age at disease onset and genotype at 3 SNPs achieved moderately high performance (AUC 0.75 - 0.78) in both the Training Set in which it was developed and a held-out Test Set. Finally, we developed a metric, the ADNC-RS, based on our logistic regression model, and demonstrated that this simple tool can identify a population of LBD individuals at very high risk for development of concomitant AD pathology.

Our findings have clinical implications. Both “proteinopathies” defining ADNC – plaques composed of amyloid-beta and neurofibrillary tangles composed of tau – are targetable with drugs in clinical trials now, and, in clinical AD, immunological approaches targeting amyloid-beta have shown enough promise to proceed to Phase III trials [5, 26]. However, within the clinical AD spectrum, the need to identify individuals ever-earlier in the course of pathophysiology [34] in order to see benefit with these therapies has created considerable problems with feasibility, not to mention potential burden to the healthcare system should any of these therapeutics attain FDA approval. These practical issues have been compounded by the fact that genetic risk scores based on AD GWAS-nominated variants achieve only very modest predictive value in the general population, where the absolute prevalence of AD is relatively low [10]. The performance of such genetics-based risk scores may be vastly improved in a population enriched for the presence of AD pathology, however [8].

Patients with primary clinical diagnoses of PD during life (and LBD at autopsy) represent exactly such an AD pathology-enriched population. Indeed, the prevalence of concomitant AD pathology in this group has been reported to range from 38% to 70%, depending on the definition of AD pathology used, and on whether clinical diagnosis of PD or primary pathological diagnoses of LBD is used [29, 33]. Our study corroborates these findings, with ~38% of PD/DLB individuals demonstrating an intermediate to high degree of ADNC, and only ~20% showing no ADNC. As a consequence, in this enriched population, the logistic regression model developed here achieves an AUC of ~0.781.

More important from a practical perspective, we use the predictors (and associated weights) identified in our model to develop a risk score for ADNC (the ADNC-RS) that can identify those PD/DLB individuals most likely to exhibit ADNC at autopsy. Indeed, in both our Training and Test Sets, those individuals with ADNC-RS in the top 20% are four times more likely to develop ADNC than LBD individuals with ADNC-RS in the bottom 40%. Because the ADNC-RS requires knowledge of only the age at disease onset and genotype at 3 AD risk SNPS, it can be easily calculated in most settings using results from a simple blood sample. Thus, the ADNC-RS developed here might serve as a screening step enriching for those PD/DLB individuals who warrant assessment for development of ADNC using more expensive modalities such as Aβ or tau imaging.

How certain can we be of our model and associated risk score? While the definitive answer to this question will lie in future studies investigating other cohorts, several aspects of our current study increase confidence. First, we nominate candidate genetic variants for inclusion in model development in an unbiased manner, starting with all loci reported to associate with risk for AD across two or more major GWAS studies. Second, we use strict criteria that are widely accepted in the field for defining ADNC. Third, and most importantly, we employ a Training Set/Test Set design in our analyses, with each group defined by consecutive genotyping of autopsy cases diagnosed with PD or DLB. Such a design guards against over-fitting, and our results confirm that we are not over-fitting the Training Set data, since performance in the Test Set is as high as in the Training Set.

Limitations of the current study should be considered alongside the previously-mentioned strengths. In particular, although our sample size of 208 neuropathologically-characterized PD/DLB cases is not small, a larger sample, across multiple centers, would be a valuable addition to the work presented here. In addition, further investigations of the cognitive consequences of ADNC in PD or DLB patients would add clinical depth to our findings. Finally, because the focus of this study was neuropathological, we defined our cohort neuropathologically, rather than using a clinical diagnosis of PD. That said, a subset analysis of the 163 individuals in our LBD cohort with a clinical diagnosis of PD yielded near-identical results. In the future, however, a clinically-defined study in a PD population, verifying the presence or absence of ADNC by imaging, could extend the current work.

In addition to the clinical implications discussed above, the biological implications of our study are also worth considering. Specifically, the genetic loci identified in our final model predicting ADNC in LBD individuals were *APOE, BIN1*, and *SORL1*. Many functions for *APOE* have been reported, but a consistent finding over many years is that the *APOE* E4 allele (included in our predictive model) encodes a form of this protein that binds Aβ less efficiently [37]. *BIN1* encodes a protein that functions in beta-secretase 1 trafficking, which in turn can impact the production of Aβ. *SORL1* encodes the sortilin-related receptor 1, which also functions in intracellular trafficking, including the sorting of APP to the retromer pathway for degradation or to the endosome-lysosome system, where APP is cleaved to generate Aβ. Collectively, the fact that our best predictive model incorporates these three genetic loci underscores the importance of Aβ production and processing in the development of ADNC among LBD individuals. In addition, direct interaction between *BIN1* and tau regulates tau phosphorylation, which may affect the development of AD pathology via a different route [20]. Interestingly, among these three genetic loci, the *SORL1* locus exerted the strongest effect in our model, with a coefficient of ~1.5 compared to ~1 for the *APOE* locus. In contrast, in the general population, among AD common genetic risk, *APOE* has by far the largest effect size.

In summary, we present our findings from a study of 208 PD/DLB cases, demonstrating that ADNC is highly prevalent in this population, and that age at disease onset and genotype at 3 SNPs is sufficient to identify a subset of LBD individuals at very high risk for development of concomitant AD pathology. The development of molecular tools such as the ADNC-RS reported here may in turn be permissive for strategies to target Aβ and tau accumulation in PD and other LBD.

## Data Availability

Data will be made available upon final publication, or by request.

## ACKNOWLEDGMENTS

We would like to acknowledge Nick Cullen and Marijan Posavi for their assistance with formulating analyses in this paper. We thank Travis Unger for technical assistance. We additionally thank our patients and their families for their generosity in contributing to this research.

This research was supported by the NIH (RO1 NS115139, P30 AG010124, U19 AG062418) and a Biomarkers Across Neurodegenerative Diseases (BAND) grant from the Michael J. Fox Foundation/Alzheimer’s Association/Weston Institute. Alice Chen-Plotkin is additionally supported by the Parker Family Chair.

